# Improving Clinical Decision-Making in Radiotherapy: A Comparative Analysis of LQ and LQL Dose Models

**DOI:** 10.1101/2025.03.05.25323402

**Authors:** Cyril Voyant, Daniel Julian, Stéphane Muraro, Véronique Bodez, Morgane Pinpin, Delphine Leschi, Rashid Oozeer, Wided Guiddi, Marie-Aimée Acquaviva, Severine Prapant, Omar Gahbiche, Noureddine Bouaouina

**Author notes:** Email addresses:* (Cyril Voyant), (Daniel Julian), (Stéphane Muraro), (Véronique Bodez), (Morgane Pinpin), (Delphine Leschi), (Rashid Oozeer), (Wided Guiddi), (Marie-Aimée Acquaviva), (Severine Prapant), (Omar Gahbiche), (Noureddine Bouaouina).

## Abstract

Radiotherapy is an essential component of cancer treatment, requiring accurate dose planning to optimize tumor control while sparing healthy tissues. This study, originating from a radiobiology workshop held during the *27*^*th*^ *Congrès National de Cancérologie et de Radiothérapie-2024* in Sousse, Tunisia, aims to investigate advanced dose modeling approaches, focusing on the Linear Quadratic (`LQ`) and Linear Quadratic Linear (`LQL`) models, to refine the calculation of biologically effective doses (`BED`) and improve treatment personalization. The workshop brought together experts in the field to discuss and evaluate the latest advancements in dose modeling, providing a comprehensive overview of current best practices and emerging trends. Using tools such as `LQL-equiv` and other `BED` calculators, we integrated patient-specific data (e.g., fractionation schedules and organ-at-risk (`OAR`) constraints) to predict outcomes such as normal tissue complication probabilities (`NTCP`). Unlike many theoretical studies, our approach embeds these models within a unified interface tailored to real clinical scenarios, enabling practitioners to simulate and adjust treatment plans based on complex, practical constraints. Through a series of clinical case studies (including treatment interruptions, palliative boosts, and reirradiation scenarios), participant responses were analyzed using the Jaccard similarity index, revealing a significant lack of consensus in treatment planning decisions (mean agreement of 25.83%). This variation illustrates the current ambiguity among clinicians regarding which model to use and how to apply it, despite access to advanced tools. This heterogeneity in decision-making could lead to divergent treatment recommendations for patients with clinically similar profiles. While the `LQ` and `LQL` models offer promising tools for personalized radiotherapy, their interpretation and implementation remain highly variable. In addition, the question of professional responsibility in dose equivalence calculations emerged as a key issue, as many departments lack clearly defined accountability frameworks. This study emphasizes the need for standardized guidelines, enhanced training programs, and decision-support systems to reduce inter-observer variability and ensure effective clinical adoption, ultimately improving patient care. The findings underscore the importance of harmonizing predictive modeling practices to achieve more consistent and effective radiotherapy outcomes.

## 1. Introduction

Modern advancements in radiotherapy and dose modeling stem from foundational developments in radiobiology. This study, originating from a workshop on advanced radiotherapy modeling, revisits key milestones to highlight the tools and concepts that have shaped contemporary clinical approaches. The workshop brought together experts in the field to discuss and evaluate the latest advancements, providing a comprehensive overview of current best practices and emerging trends. While the `LQ` and `LQL` models are well established, their clinical application remains highly variable and underreported. This variability may result in significantly different therapeutic strategies for patients facing comparable clinical conditions. This paper quantifies, for the first time to our knowledge, the variability in clinical interpretation and use of these models across different professionals and scenarios. Quantifying this inconsistency is critical to guide training priorities and support the development of harmonized clinical protocols.

### 1.1. Text Mining In Context

Artificial intelligence, and in particular `AI` transformer-based models, are increasingly used in medicine to extract insights from unstructured biomedical literature. In radiotherapy and radiobiology, where concepts like dose, tissue response, and biological modeling are complex and evolving, `AI`-driven text mining enables scalable identification of patterns across large publication corpora. This study applies an `AI` transformer model (`all-MiniLM-L6-v2`) to semantically filter (Natural Language Processing, *i*.*e*. `NLP`) and score scientific articles related to radiobiological metrics. Publications were sourced from `Scopus` and enriched using `CrossRef` and `OpenAlex` to support a multi-dimensional analysis. The resulting dataset includes 1,118 publications spanning 2000 to 2024. Among the documents with specified types, 59.7% are journal articles, the rest is related to other types of publications, including reviews, conference papers, and book chapters. The open access rate stands at 25.0%, and the average citation count is 25.2 per paper. Temporal analysis reveals a statistically significant linear growth in publication volume over time (*slope* = 0.7/year, *p* = 0.001, *R*^2^ = 0.42), with no indication of exponential acceleration, suggesting a steady and sustained interest in radiobiological evaluation. Notably, only 26.7% of the publications explicitly reference mathematical dose modeling (*e*.*g*., `EQD2`, `BED`, or the `LQ` model), reflecting a clinical dominance in the literature. In terms of productivity, Jones B., Dale R.G., and Mavroidis P. are the most prolific authors. However, the most influential based on cumulative citations are Jones B. (908), Joiner M.C. (744), and Marples B. (709). Geographically, institutions from the United States (45.8%), Japan (25.3%), and France (16.0%) are the most represented. Leading journals include the *International Journal of Radiation Oncology Biology Physics* and *Medical Physics*, published primarily by `John Wiley` and `Elsevier`, underscoring their central role in disseminating high-impact radiobiological research.

### 1.2. Historical Background in Radiobiology

Radiotherapy has evolved significantly since Lea and Catcheside’s work on indirect radiation effects [1]. The Linear Quadratic (`LQ`) model, pivotal in the 1980s, facilitated fractionation modeling [2] and became a cornerstone for dose calculation due to its simplicity and broad applicability. However, its limitations at high doses, such as overestimating cell survival, led to the development of refined models. Lyman’s dose-volume histograms improved the prediction of tissue complications [3], providing a more accurate assessment of the risk to healthy tissues. Fowler’s Biologically Effective Dose (`BED`) standardized dose equivalence across different fractionation schemes [4], making it easier to compare different treatment protocols. Dale further enhanced the model by incorporating tumor repopulation effects [5], which are crucial for accurately predicting treatment outcomes. To address the high-dose limitations of the `LQ` model, Astrahan proposed the Linear Quadratic Linear (`LQL`) model [6], which better accounts for cell survival at higher doses. This model is particularly useful in scenarios involving high-dose treatments, such as stereotactic radiosurgery. Niemierko’s introduction of the Equivalent Uniform Dose (`EUD`) provided a framework for handling dose inhomogeneities [7], allowing for more precise treatment planning. Recent advancements have integrated Relative Biological Effectiveness (`RBE`), particularly for ion and proton therapies, further refining dose calculation accuracy [8, 9]. These models consider the varying biological effects of different types of radiation, improving the precision of dose calculations. Despite these advancements, the clinical application of these models remains inconsistent. This study aims to evaluate the `LQ` and `LQL` frameworks in the context of modern radiotherapy, highlighting their strengths, limitations, and potential for standardization. Numerous studies and guidelines have recently addressed the challenges of dose calculation and standardization in radiotherapy. As an example, Hardcastle et al. conducted a multi-centre study assessing variability in cumulative dose reporting for reirradiation scenarios [10]. Despite using similar datasets and objectives, their results demonstrated large variations in calculated doses across institutions, underscoring the impact of differences in methodology and interpretation. These findings strongly support the need for standardized workflows when applying dose models in complex clinical contexts. Similarly, the Royal College of Radiologists has provided comprehensive national guidance for managing unscheduled treatment interruptions [11]. Their fourth edition outlines radiobiological calculations and proposes concrete solutions to ensure safe and effective continuity of treatment, including model-based compensation methods. These examples illustrate the increasing awareness of inconsistencies in clinical application of well-established models such as `LQ` and `LQL`. The present study aims to contribute to this collective effort by highlighting not only the theoretical underpinnings of these models, but also their varied use. In doing so, we hope to support future harmonization of model application and facilitate the integration of biologically informed decision-making into routine practice.

### 1.3. Legislation and Responsibility

Modern radiotherapy is governed by stringent guidelines to ensure safety and efficacy. Regulatory frameworks, such as EU Regulation 2017/745 and French decrees (e.g., R. 4127-8 and R. 4351-1), mandate accurate dose calculation and strict adherence to organ-at-risk (`OAR`) constraints. These regulations require continuous monitoring of patient outcomes and adaptive strategies to address treatment interruptions, evolving patient conditions, and technological advancements. Practitioners must balance clinical responsibilities with legal requirements, as outlined in international safety standards [12]. For a detailed overview of the roles and responsibilities of key stakeholders, see Figure 1 and Table 1. This figure illustrates the chain of responsibility and coordination in radiotherapy dose calculation workflows, based on EU and French regulatory standards. While some roles are jurisdiction-specific, the structure reflects widely applicable clinical functions. The “Radiology Technician” (French: Manipulateur en électroradiologie médicale) refers to the role often known internationally as Radiation Technologist or RTT (not to be confused with Radiation Oncologists or Dosimetrists). All clinical actions involving dose calculations remain under the authority and validation of the Medical Physicist and Radiation Oncologist. To minimize risks associated with software defects or misuse, healthcare facilities should implement robust quality assurance programs. These programs should include regular software audits, comprehensive training for staff, and clear protocols for dose calculation and verification. Such measures not only ensure compliance with legal standards but also enhance patient safety and treatment efficacy. While the European, and more specifically French, framework was used as a reference in this discussion, the conclusions regarding responsibility in radiobiological decision-making are universal in nature and may be considered applicable, at least in part, across various international healthcare systems.

**Table 1:** Comparison of Clinical and Legal Responsibilities in Cases of Software Defects and Misuse in Radiotherapy.

| Case Study | Defective Software: Patient dies or suffers irreversible harm due to a software defect causing incorrect dose calculations. | Misuse of Software: Patient dies or suffers irreversible harm due to a user error in dose calculation. |
| --- | --- | --- |
| Primary Responsibility | Developer: Responsible for testing and safety compliance (e.g., *EU Reg. 2017/745*). Negligence liability if a defect caused harm. | Physicist/Oncologist: Responsible for accurate data entry and following protocols. Liability if misuse caused harm. |
| Secondary Responsibility | IT Department: Accountable if risk monitoring was insufficient to catch defects. | Healthcare Facility Director: Responsible if training or guidance was inadequate. Liability for insufficient protocols or oversight. |
| Shared/Indirect Responsibility | Facility Director: Liable if the software was uncertified or improperly vetted for clinical use. | Developer: Some responsibility if unclear interface contributed to error. Must ensure software is intuitive and provides clear instructions. |
| Legal Implications | - Negligence for the developer or IT if defect was undetected.<br>- Regulatory breach if uncertified software was used, per EU standards. | - Malpractice liability for the healthcare professional if error stemmed from user negligence.<br>- Institutional liability if misuse was due to inadequate training. |

**Figure 1:**
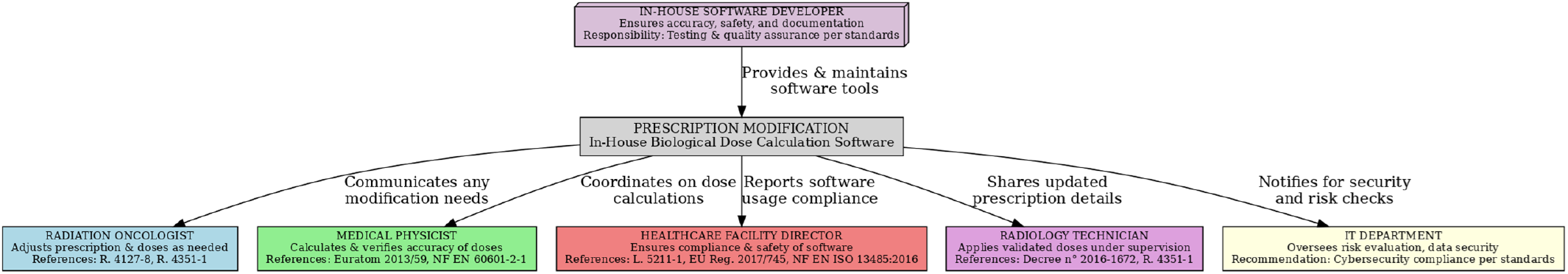
Overview of Legal Responsibilities in Radiotherapy Dose Calculation Workflow (EU/French Example). Note that the role of “Radiology Technician” Corresponds to Radiation Technologist/RTT)

### 1.4. The Importance of BED and NTCP

The Biologically Effective Dose (`BED`) has become a key element for quantifying the biological impact of different fractionation schemes, enabling direct comparison of diverse protocols [13]. By accounting for dose per fraction, total dose, and tissue-specific parameters, `BED` provides a unified metric to evaluate and optimize treatment plans. For example, it allows clinicians to compare conventional fractionation (e.g., 2 Gy per fraction) with hypofractionated regimens (e.g., 5 Gy per fraction) while maintaining equivalent biological effectiveness. Similarly, the Normal Tissue Complication Probability (`NTCP`) metric facilitates risk estimation for organ damage, guiding personalized treatment planning. `NTCP` models integrate dose-volume histograms and radiobiological parameters to predict the likelihood of complications, such as radiation-induced pneumonitis or fibrosis. However, their accuracy depends on the quality of the underlying data and the applicability of the model to specific patient populations. Together, `BED` and `NTCP` offer a robust framework to optimize treatment outcomes and manage toxicity. Despite their widespread use, challenges remain, including inter-patient variability and the need for standardized implementation across clinical centers. This study aims to address these challenges by evaluating the consistency and applicability of these metrics in concrete scenarios.

### 1.5. Objectives

This study, originating from a workshop on advanced radiotherapy modeling, investigates the integration of sophisticated dose models, particularly the Linear Quadratic (`LQ`) and Linear Quadratic Linear (`LQL`) frameworks, in radiotherapy planning. One of its key aims is to validate the applicability of Biologically Effective Dose (`BED`) and Normal Tissue Complication Probability (`NTCP`) metrics across diverse clinical scenarios. Additionally, it evaluates the consistency of participant responses by employing the Jaccard similarity index [14], revealing potential variability in treatment planning decisions. The study also examines the practical implementation of `LQ` and `LQL` models through a series of case studies, including treatment interruptions and reirradiation cases, to assess their relevance. While the theoretical foundations of these models are well established, their clinical application remains highly heterogeneous. This work highlights the gap between the availability of modeling tools and their real-world use, especially in scenarios where clinicians face uncertainties regarding model selection and implementation. To address this challenge, the study incorporates an integrated software interface (LQL-equiv) that consolidates model computations, clinical constraints, and patient-specific parameters, allowing for a realistic assessment of model usability in day-to-day clinical workflows. The observed variability among responses illustrates the urgent need for clearer decision-support mechanisms and harmonized protocols.Furthermore, the study explores an often-overlooked issue: the attribution of responsibility in dose-equivalence calculations within radiotherapy departments. Through real-world case scenarios, we aim to identify key stakeholders (physicians, physicists, software developers, institutions) involved in the process, and highlight the absence of standardized accountability frameworks. This raises important questions regarding safety, training, and legal liability in modern radiotherapy practices. Finally, it advocates for the development of standardized guidelines to facilitate the adoption of advanced dose modeling, aiming to minimize inter-observer variability and enhance clinical decision-making. By addressing these aspects, the study aims to document and quantify the existing gap between theoretical radiobiological models and their clinical application. This work represents an essential preliminary step toward harmonizing practices and developing more standardized, evidence-based approaches to dose modeling in radiotherapy.

## 2. Methods

`LQL-equiv` is an open-source tool for dose-equivalence modeling. Built on the Linear Quadratic Linear (`LQL`) framework, `LQL-equiv` addresses the limitations of traditional `LQ` models, particularly for extreme fractionations. The tool was selected for this study due to its integration of advanced radiobiological models (`LQ`, `LQL`, and `BED`), its adoption in over 20 countries, and its ability to comprehensively simulate complex clinical scenarios. While `LQL-equiv` is a research-oriented tool without clinical certification (a limitation shared by many dose modeling software), it offers one of the most comprehensive integrations of radiobiological models available. This experimental nature reinforces the importance of conducting formal clinical validation before adopting such tools into routine practice. `LQL-equiv` supports `LQ, LQL, BED, NTCP` calculations across a wide range of tumors and organs, and uniquely includes parameters such as kick-off time, repopulation, and dose constraints. Its built-in optimization engine (not found in other clinical or academic tools to our knowledge) enables users to simulate and refine treatment plans using biologically driven targets. This allows clinicians not only to test theoretical models, but to calibrate and validate them in realistic scenarios, thereby enhancing the clinical relevance and decision-making potential of advanced dose modeling. The setup realistically mirrors clinical conditions where multiple dose models coexist, forcing practitioners to make context-sensitive therapeutic choices.

### 2.1. Workflow of LQL-equiv

Figure 2 outlines the workflow of `LQL` *−* `equiv`. Data acquisition consists of inputting patient-specific data, including total dose, dose per fraction, pause days, and fractionation schedule. The Linear-Quadratic Linear (`LQL`) model is then applied to compute the Biologically Effective Dose (`BED`):

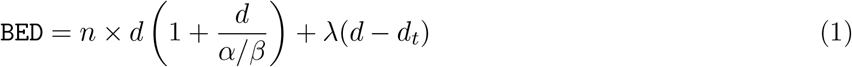

where *n* is the number of fractions, *d* is the dose per fraction, *α/β* is the tissue-specific parameter, *λ* accounts for treatment breaks, and *d*_*t*_ is the threshold dose. For the target volume (`TV`), tumor repopulation effects are considered:

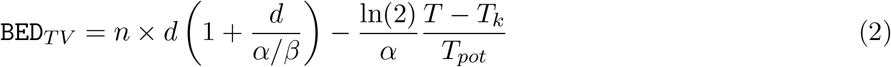

where *T*_*k*_ represents the kick-off time for tumor repopulation, *T*_*pot*_ is the potential doubling time, and *T* is the total treatment duration. For the organ at risk (`OAR`), the `BED` calculation accounts for repair capacity and dose constraints:

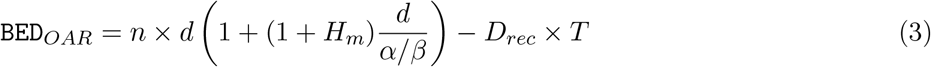

where *H*_*m*_ represents the repair factor, *D*_*rec*_ is the recommended maximum dose, and *T* is the treatment duration. To enable comparisons across fractionation schemes, the Equivalent Dose in 2 Gy Fractions (`Eq2Gy`) is computed as:

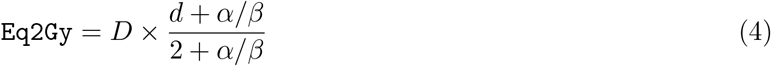

**Figure 2:**
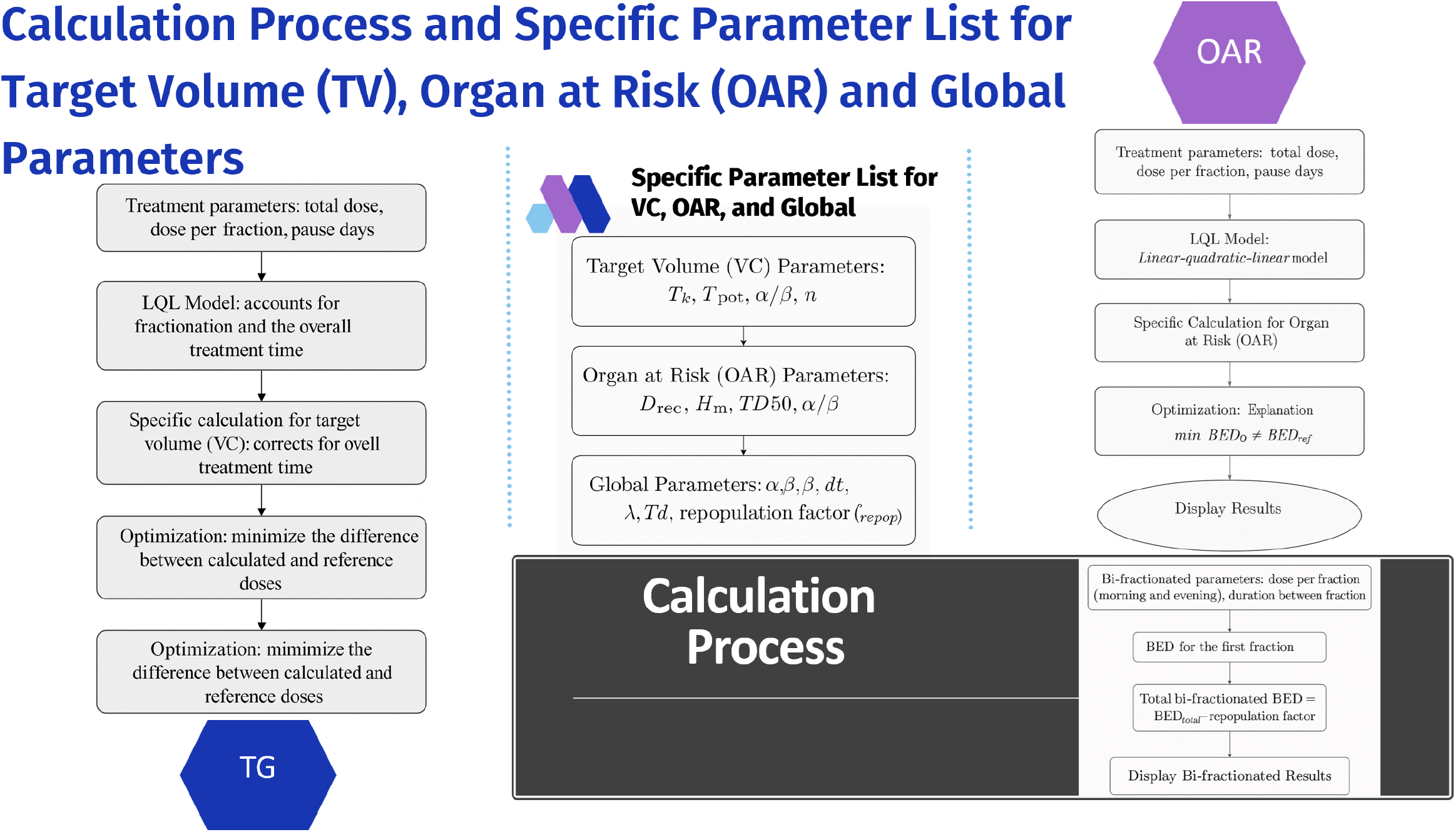
Workflow of `LQL-equiv`: From Clinical Parameters to Dose Validation. Key Steps and Required Inputs for Target Volumes, Organs at Risk, and Global Factors.

Optimization is performed to adjust treatment parameters and minimize the difference between the calculated `BED` and the reference `BED`:

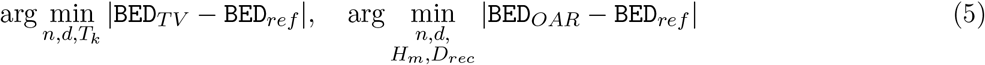

For bi-fractionated treatments involving two daily dose fractions, the total `BED` is computed as:

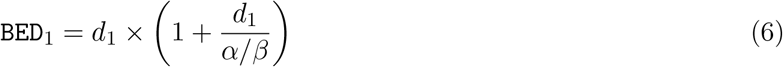

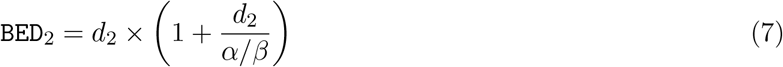

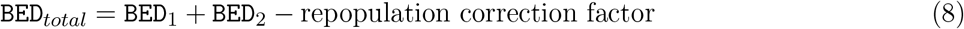

Finally, the workflow generates optimized treatment plans, including dose adjustments and `NTCP` estimates. This framework integrates tumor-specific and `OAR` constraints into `BED` and `Eq2Gy` calculations, optimizing treatment personalization and minimizing complications. The use of repopulation correction and fractionation adjustments allows for precise radiotherapy planning.

### 2.2. Participant Evaluation and Metrics

Participants in this study were attendees of the *27*^*th*^ *Congrès National de Cancérologie et de Radiothérapie-2024*, held in Sousse, Tunisia, on November 22–23, 2024. This event gathered over 1,000 registrants, with approximately 20% attending in person and the rest participating virtually. The diversity of attendees, including radiation oncologists, medical physicists, and radiotherapy technicians, enhanced the analysis by incorporating a broad range of perspectives and expertise. During the workshop, participants were divided into heterogeneous groups of five, each comprising a mix of physicians, physicists, and technicians. This structure was designed to foster interdisciplinary collaboration and encourage knowledge sharing, promoting a deeper understanding of the challenges and solutions in radiotherapy dose optimization. Each problem (four in total) was first introduced by the facilitators (C. Voyant and D. Julian), providing the necessary context and theoretical background. Groups were then given time to discuss and respond to each case using a provided paper-based booklet. This strategy enabled us to isolate individual decision-making patterns from collective group influence, revealing nuanced inter-practitioner variability. At the end of the session, each participant submitted their completed paper booklet. The data were then digitized and analyzed by the authors using a homemade survey platform built in MATLAB. This platform allowed for anonymized and structured entry of the handwritten responses to compute descriptive statistics and similarity indices. Each group had access to a dedicated computer equipped with `LQL-equiv` and other dose calculation tools^1^. These tools were selected to provide a comprehensive comparison of different dose calculation methodologies, enabling participants to explore their respective strengths and limitations. Participants alternated in using the software, fostering a dynamic and interactive approach to problem-solving. Discussions within the groups allowed for collaborative decision-making and real-time exploration of complex cases. This interactive format ensured that all participants, regardless of their background, could contribute to the decision-making process. Furthermore, the collaborative environment facilitated the exchange of best practices and reinforced the importance of interdisciplinary teamwork in optimizing radiotherapy treatments.

### 2.3. Data Collection and Analysis

The clinical cases used in this study were pre-defined to evaluate participant decision-making in scenarios such as treatment interruptions, palliative boosts, and reirradiation (details in Appendix A). Each case included specific dosimetric constraints (e.g., maximum spinal cord dose, mean cardiac dose) and required participants to propose adjusted treatment plans. Responses were collected via a homemade survey platform (using MATLAB), ensuring independent and anonymized participation. The Jaccard similarity index (see Appendix B for more information) was used to quantify agreement among participants, with additional summary statistics (e.g., response rates, most frequent adjustments) providing further insights into decision-making patterns. While there are many mathematical tools available to perform this operation, we aimed to keep it very simple and as objective as possible, knowing that the goal was solely to check if the responses (to one decimal place) were homogeneous or not. The global response rate for all answers (3 per clinical case as described in Appendix A) is close to 70%. This suggests that some participants either did not have enough time or were unsure of the method to use. To further characterize response variability, standard descriptive statistics including standard deviation range were computed for each clinical scenario. Additionally, `Kruskal-Wallis` tests were applied to the proposed dose per fraction values across cases to assess whether inter-case differences were statistically significant. This non-parametric method was selected due to the non-normal distribution of the response data. All analyses were conducted using `MATLAB` software. For further details on `LQL-equiv`, including download links and technical documentation, visit:

✓ https://github.com/cyrilvoyant/LQ-Equiv
✓ https://cloud.minesparis.psl.eu/index.php/s/dzoXsjaD9MCc5aw/download

For additional information, see Voyant et al. [15, 16].

## 3. Results

The analysis of participant responses demonstrated diverse strategies for addressing clinical challenges, reflecting the variability in decision-making even among experienced practitioners. During the congress, the practical relevance and variability of radiobiological calculations were widely debated, with many clinicians expressing uncertainty about how to consistently apply the models in daily practice. This study reflects those concerns and provides a first step toward clarifying how such models are currently interpreted in clinical settings. To illustrate these findings, case studies are detailed, beginning with an interrupted treatment scenario. These examples highlight participant reasoning and adjustments, revealing common trends and deviations. Full details of the clinical scenario and associated question are provided in Appendix A. For reference, a sample solution calculated by the authors using `LQL-Equiv` is also included.

### 3.1. Case Study 1: Treatment Interruption in Curative Radiotherapy

This case involved a patient undergoing curative radiotherapy who experienced a one-week treatment interruption due to radiation-induced skin toxicity. The interruption occurred after two weeks of treatment, with 10 fractions already delivered. Participants were tasked with determining the optimal therapeutic plan to resume treatment while adhering to strict dosimetric constraints. The clinical and dosimetric challenge required maintaining the original number of planned fractions while ensuring the safety of critical organs-at-risk (`OAR`). Specific criteria included:

✓ Maximum spinal cord dose (`PRV`): 46 Gy.
✓ Mean cardiac dose (`EUD`): Below 7 Gy.
✓ Maximum bowel dose (`V50`): Under 10%.

Participants evaluated potential acute and late treatment effects, ensuring that the Normal Tissue Complication Probability (`NTCP`) remained below 5%. This scenario required individualized dose adjustments for three anatomical regions: head and neck (`ORL`), breast, and bladder. Adjustments to the prescribed fraction doses were recorded to ensure compliance with clinical and dosimetric constraints for each region. The analysis revealed significant variability in participant responses, with proposed fraction doses ranging from 1.8 Gy to 2.5 Gy for the head and neck region, 2.0 Gy to 2.4 Gy for the breast, and 1.9 Gy to 2.3 Gy for the bladder. This variability underscores the challenges of standardizing dose adjustments in complex clinical scenarios, even when using advanced tools like `LQL-Equiv`. This highlights a need for clearer clinical recommendations to guide practitioners in scenarios where dose compensation is required. The most frequent adjustments (mode) were 2.2 Gy for the head and neck, 2.1 Gy for the breast, and 2.1 Gy for the bladder, indicating a tendency toward moderate dose escalation to compensate for the interruption.

### 3.2. Case Study 2: Palliative Boost for Cerebellar Metastasis

This case involved a patient with a symptomatic cerebellar metastasis from breast cancer, undergoing palliative radiotherapy. The standard protocol comprised whole-brain irradiation delivering 30 Gy in 10 fractions. Due to severe symptoms, a targeted dose escalation was required for the cerebellar lesion to improve local tumor control and symptom relief while adhering to strict dosimetric constraints. The clinical objectives were:

✓ Deliver at least 45 Gy (`Eq2Gy`) to the metastasis.
✓ Limit the maximum brain dose to 50 Gy (`Eq2Gy`).
✓ Protect vision-related `OAR`, keeping doses below 45 Gy (`Eq2Gy`).

Participants evaluated two therapeutic approaches, Sequential Boost (administered after whole-brain irradiation, requiring specification of the number of fractions and dose per fraction) and simultaneous Integrated Boost (`SIB`; integrated into whole-brain treatment, requiring only the dose per fraction to be recorded). They were tasked with selecting one of the approaches and providing a global decision. For both strategies, adherence to clinical objectives and prioritization of shorter treatment schedules, where feasible, were emphasized.

### 3.3. Case Study 3: Treatment Interruption and Resumption

This case involved a metastatic lung cancer patient undergoing radiotherapy targeting the L2-L3 vertebrae. The initial treatment plan prescribed 30 Gy over 10 fractions. After completing 5 fractions, a two-week interruption occurred due to hospitalization. Participants were tasked with proposing a therapeutic plan to resume treatment while maintaining the same biological effectiveness as initially planned. Two resumption strategies were considered:

✓ Maintaining the remaining 5 fractions.
✓ Modifying the number of remaining fractions.

In both strategies, participants had to ensure that the maximum spinal cord dose remained below 34 Gy (physical dose). For each approach, participants calculated the equivalent biological dose for various fractionation schemes and proposed an updated plan aligned with these constraints. The clinical challenge involved recalculating fractionation to achieve the original therapeutic objectives. For each strategy, the dose per fraction was specified, and participants provided a global decision on whether to maintain or modify the number of fractions.

### 3.4. Case Study 4: Spinal Cord Dose Limitation in Reirradiation

This scenario involved a patient previously treated 10 years ago for breast cancer, receiving a maximum dose of 23 Gy to the spinal cord over 23 fractions (26 Gy to the planning risk volume, `PRV`, of the spinal cord +5 mm) on a volume of 0.03 cc. The patient now requires reirradiation targeting the T3 vertebral body with a proposed prescription of 5 fractions of 7 Gy each. The clinical challenge was to limit spinal cord toxicity while ensuring effective tumor control. Participants calculated dose equivalence using the 2 Gy equivalent dose (`Eq2Gy`) or Biologically Effective Dose (`BED`) models, accounting for accumulated doses from prior treatments. Constraints from the International Spine Radiosurgery Consortium Guidelines were strictly applied:

✓ Maximum dose (`Dmax`) to the spinal cord: *<* 28 Gy (0.03 cc).
✓ Volume receiving *>* 22 Gy (`V22`Gy): *<* 0.35 cc.

Participants were encouraged to use `LQ` or `LQL` frameworks to evaluate fractionation schemes and ensure compliance with dose constraints. They determined the dose per fraction for spinal cord protection under three fractionation schemes (5, 6, or 7 fractions). Each option required specifying the dose per fraction to balance therapeutic effectiveness and spinal cord safety.

### 3.5. Discussion

The findings of this study highlight the potential of advanced dose modeling in radiotherapy treatment planning. However, the analysis of participant responses to clinical case studies revealed significant variability in the proposed solutions. As shown in Table 2, the Jaccard similarity index (`J`) indicated a mean agreement of only 25.83% among participants. To the best of our knowledge, no other study has measured consensus or clinical heterogeneity on these models via case studies and `J` makes it possible to objectively quantify this variability, which is in itself an original contribution. To further quantify the dispersion of participant responses, the standard deviation (`SD`) of proposed fraction doses was computed for each anatomical site. The resulting `SD` values ranged from 0.14 Gy to 0.31 Gy across all case studies, indicating moderate variability even within the most frequently chosen dose levels. This statistical spread reinforces the interpretation of clinical heterogeneity beyond `J` alone. `Kruskal-Wallis` tests comparing dose distributions across participant responses revealed statistically significant differences in Case 1 for all anatomical regions: head and neck (*p* = 0.018), breast (*p* = 0.032), and bladder (*p* = 0.027). Similar significance levels (*p <* 0.05) were observed across the remaining three case studies, indicating that the dose variability between scenarios is statistically meaningful.

**Table 2:**
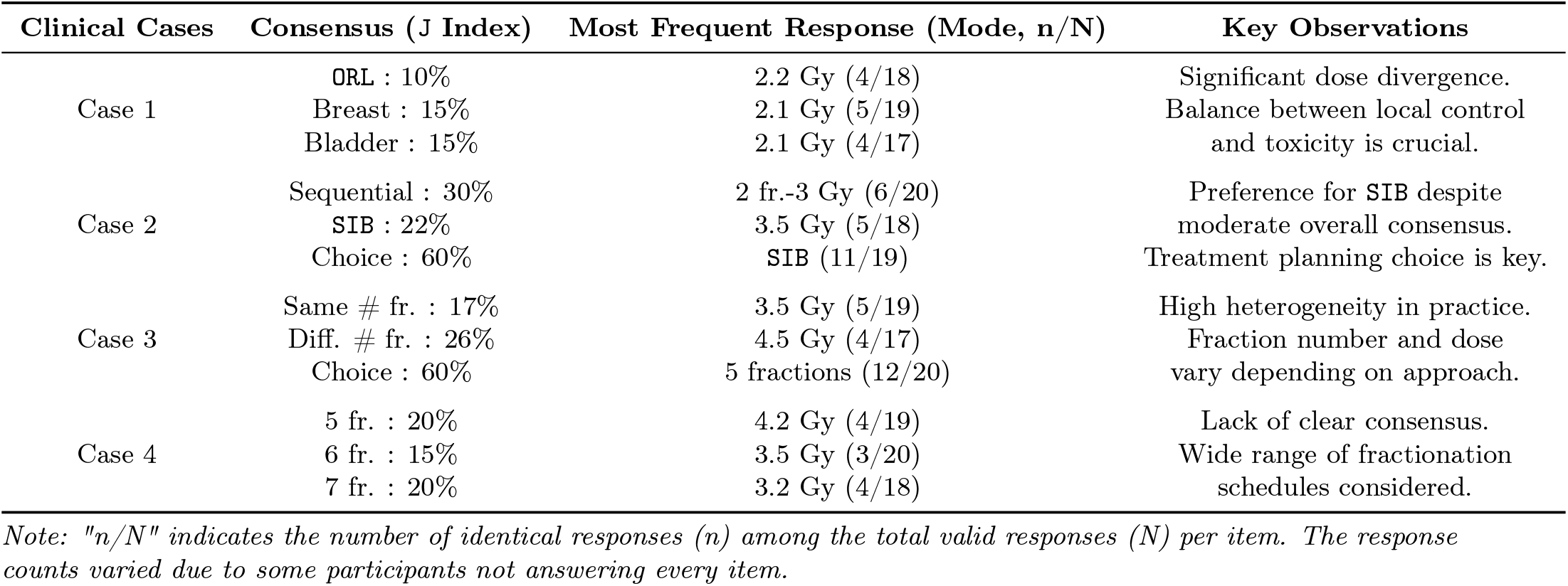
Analysis of Response Variability Across Case Studies Using Jaccard Index (J) and Mode with Frequencies (n/N)

This low concordance highlights several crucial points. Firstly, it reveals that even when using dose-equivalence calculation tools such as `LQL-equiv` and other available `BED` calculators, practitioners arrive at divergent therapeutic conclusions. This divergence persists even when participants use the same tool, suggesting that the interpretation of results and their clinical application vary considerably. The observed differences are sometimes disconcerting, underscoring the inherent complexity in applying radiobiological models in real clinical contexts. Analysis of Table 2 allows us to further specify this observation:

✓ Case 1 (Treatment Interruption): The low concordance (10% for `ORL`, 15% for Breast and Bladder) associated with similar proposed doses (2.1 Gy and 2.2 Gy) indicates uncertainty regarding the optimal strategy for compensating for the interruption. Although the doses are numerically close, the biological impact of this slight difference, particularly in terms of `BED`, could be significant. This suggests that even small variations in fraction doses can generate debate regarding the benefit/risk balance.
✓ Case 2 (Palliative Boost for Cerebellar Metastasis): A marked preference (60% of choices) for the use of `SIB` is observed, despite moderate consensus on specific parameters (30% for sequential fractionation and 22% for `SIB`). This preference for `SIB` could reflect a desire for treatment simplification or a perception of better clinical efficacy, although the data do not allow us to confirm this.
✓ Case 3 (Treatment Interruption and Resumption): This case presents high heterogeneity in practice, with a distribution of choices between maintaining the number of fractions (17%) and modifying it (26%), and an overall preference for 5 fractions (60% of choices). The associated doses (3.5 Gy and 4.5 Gy) also demonstrate significant variability. This heterogeneity underscores the difficulty of establishing a standardized protocol for managing treatment interruptions and the need for an individualized approach.
✓ Case 4 (Spinal Cord Dose Limitation in Re-irradiation): The lack of clear consensus (20% for 5 and 7 fractions, 15% for 6 fractions) and the dispersion of proposed doses (between 3.2 Gy and 4.2 Gy) reflect the complexity of re-irradiation, particularly due to strict dose constraints on the spinal cord. Calculating `Eq2Gy` or `BED` in this context is crucial, but the results show that even with these tools, interpretation and clinical decisions diverge.

Several factors can explain this variability. One key issue is the heterogeneous training received by practitioners regarding the use and limitations of `LQ` and `LQL` models. While these models are well documented in the literature, their translation into clinical routine remains inconsistent. Additionally, the lack of standard guidelines on their application in specific scenarios, such as treatment interruptions or reirradiation, forces clinicians to rely on individual experience rather than established protocols. Furthermore, although digital tools such as `LQL-equiv` provide objective calculations, their interpretation remains subjective, particularly in complex cases where multiple fractionation strategies are feasible. The clinical impact of this variability is not trivial. A difference of 0.2 to 0.5 Gy per fraction, though seemingly small, can significantly alter the total biological dose (`BED`) received by the tumor and organs at risk. In the context of reirradiation, such discrepancies can lead to suboptimal tumor control if the dose is underestimated, or to severe complications if it is overestimated. This highlights an urgent need for standardized reference values and decision-support tools that can guide clinicians towards more consistent and reproducible treatment planning. Our findings align with recent work by Hardcastle et al. [10], who demonstrated significant inter-centre variability in the technical assessment of cumulative doses during reirradiation. While their focus was on differences in dose accumulation workflows using physical DICOM data and deformable registration tools, our study complements this by revealing the variability in clinical decision-making using the same radiobiological models. Together, these studies underscore that inconsistency exists not only in computational methods but also in interpretive practices, even when similar theoretical frameworks are applied. Furthermore, our results echo some of the concerns addressed in the latest Royal College of Radiologists’ guidelines on the management of unscheduled treatment interruptions [11], which advocate for structured use of BED-based compensation calculations. However, our data suggest that these guidelines, although comprehensive, are not uniformly adopted or interpreted in practice. This highlights the persistent gap between published recommendations and clinical implementation, particularly when decisions must be made under uncertainty. In contrast to both documents, which offer either technical or normative perspectives, the present study contributes a practical, observational view on how practitioners engage with these models across different clinical scenarios. This triangulated evidence (from guidelines, technical analysis, and clinical observation) collectively points to the need for more integrated, user-friendly tools and clearer clinical protocols to support the reliable application of radiobiological modeling in routine care.

## 4. Conclusion

This study explored the integration of advanced dose modeling, specifically the `LQ` and `LQL` models, into clinical radiotherapy planning. The analysis of participant responses across diverse case studies revealed a concerning lack of agreement in treatment planning decisions, despite the availability of tools like `LQL-equiv` and other `BED` calculators. This underscores that while advanced modeling offers the potential for personalized and optimized treatments, the interpretation of model outputs and their translation into clinical practice remain highly variable. Therefore, successful integration of these models requires a strong emphasis on standardization. We recommend the development of comprehensive guidelines, standardized training programs focusing on both theoretical understanding and practical application, and exploration of AI-based decision support systems to minimize inter-observer variability and ensure consistent, effective clinical adoption of advanced dose modeling in radiotherapy. In addition, this paper presents clinical cases frequently encountered in radiotherapy departments, along with solutions provided using `LQL-equiv` and its developers. The most probable responses from the sample of clinicians present at the conference are also documented. This allows each radiotherapy team to compare its practice with these results, facilitating reflection on clinical decision-making and potential improvements in treatment planning. To move towards standardization, the next step should be the creation of a practical guide on the use of `LQ` and `LQL` models in clinical settings, incorporating best practices derived from this study. Additionally, a working group could be established to foster collaboration between institutions, ensuring a harmonized approach to dose equivalence calculations. Finally, future developments should aim at integrating these models into clinically validated digital tools that facilitate decision-making and minimize subjective interpretation. Furthermore, this study highlights the critical need to define roles and responsibilities in dose-equivalence decision-making. Currently, the attribution of clinical, technical, or legal responsibility remains unclear in many institutions, potentially compromising safety and accountability. Unlike previous theoretical contributions, this study is grounded in clinical variability and provides concrete, actionable insights into current practice gaps.

Given these findings, it is urgent to repeat such workshops to guide practitioners and systematically compare practices. Establishing regular assessments would help pave the way toward the harmonization of equivalent dose calculations in the medium term. It is unacceptable that tumor control and radiation-induced effects depend on the treatment center or the individual performing the calculations, especially when technical capabilities are equivalent. Ensuring consistency in radiotherapy protocols should be a priority to guarantee optimal patient care across different institutions.

## Data Availability

All data produced in the present study are available upon reasonable request to the authors

## Acknowledgements

The authors gratefully acknowledge `Elsevier` and the `Scopus` platform for providing structured access to bibliographic and abstract metadata. Their `API` services were essential in supporting the large-scale text mining and semantic enrichment pipeline implemented in this study. Such infrastructure enables transparent, reproducible, and scalable research in scientific knowledge mapping. We would like also to express our sincere gratitude to the organizers of the *27*^*th*^ *Congrès National de Cancérologie et de Radiothérapie-2024*, and to the radiotherapy and administrative teams at Ibn Khaldoun Center (represented by M. Mohamed DAOUAS). We are deeply grateful for the organizers dedication in creating such a stimulating and informative event.

## Ethics approval

No ethical approval was required for this study, as no patient data was used. The simulations were based solely on fictitious case studies.

## Data availability Statement

The data used in this study is available upon reasonable request to the corresponding author, within the limits of feasibility.

## Appendix A. Detailed Clinical Cases

The clinical cases presented in this section illustrate the challenges posed by the radiotherapy management of patients with brain metastases. We will explore the different therapeutic strategies considered and the decision-making criteria that guided the choice of treatment. Figures A1 and A2 provide visual representations of the cases and the proposed solutions.

## Appendix B. Algorithm for Jaccard Similarity (J) and Mode Determination

The evaluation of participant responses was guided by two complementary metrics: the Jaccard similarity index and the mode. The Jaccard index quantifies the consistency of responses by calculating the proportion of shared elements between pairs of responses relative to the total. Meanwhile, the mode identifies the most frequent response, shedding light on collective tendencies in decision-making. Algorithm 1 outlines the process for computing both metrics. The Jaccard similarity measures agreement, while the mode highlights the dominant adjustment strategy. Together, they provide a comprehensive picture of participant behavior.

### Algorithm 1

Jaccard Similarity and Mode Calculation

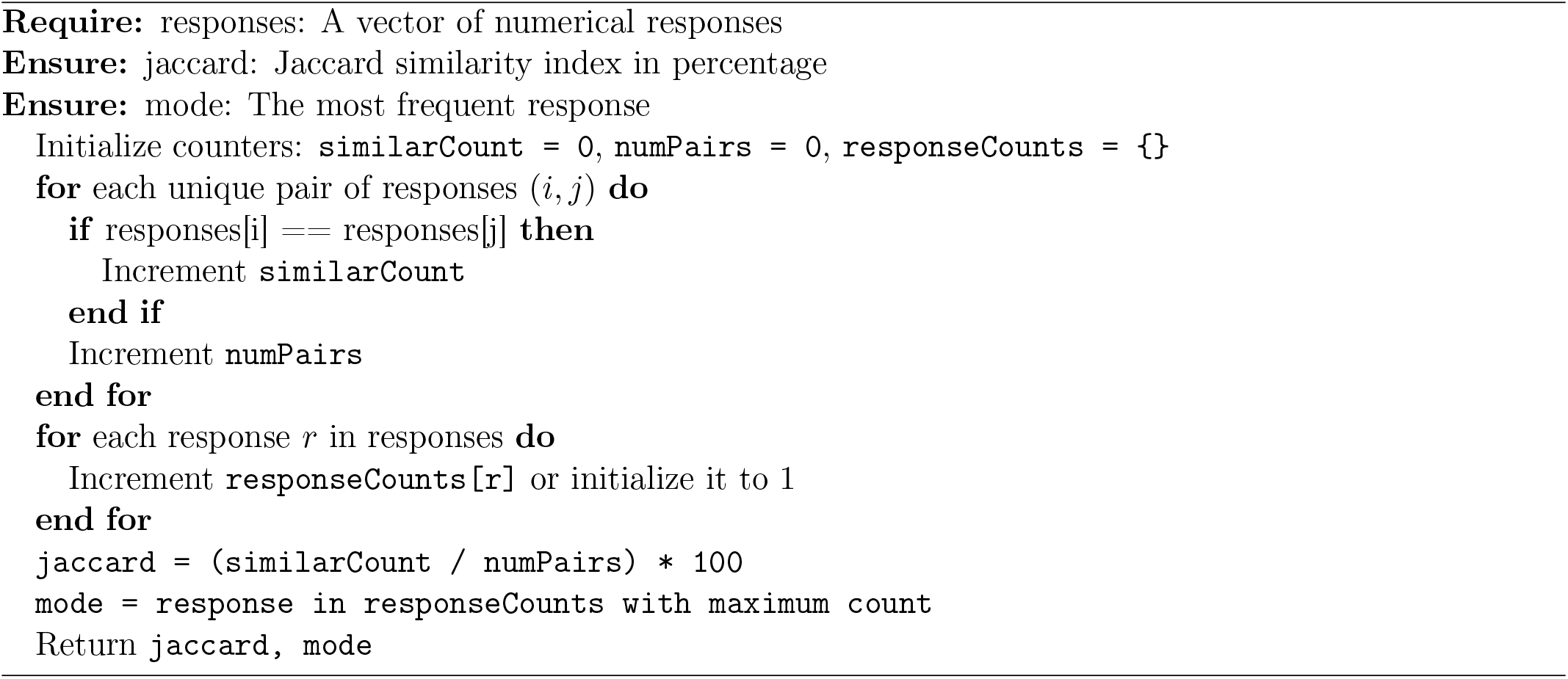

The results of these calculations reflect not only the degree of agreement among participants but also the most prevalent adjustment strategy. For example, in the case of interrupted treatments (case 3), the mode revealed that most participants preferred to maintain the duration of treatment while favoring low dose levels per fraction. Such insights are essential for understanding collective decision-making and optimizing future training efforts.

**Figure A1:**
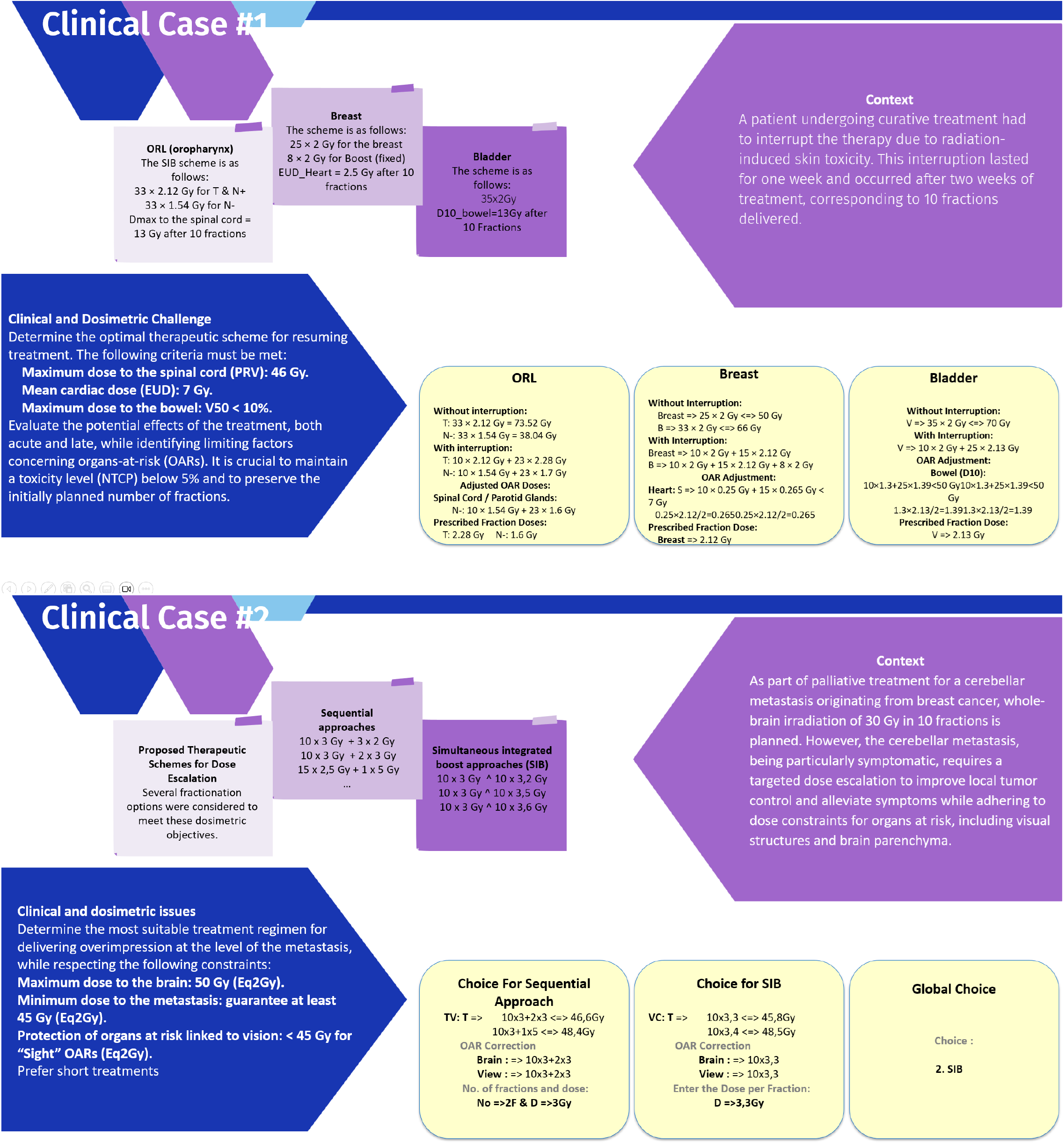
Illustration of Case Studies 1–2: Scenarios and `LQL-equiv`-Based Suggested Solutions. Case 1 – Collected dose per fraction: ORL = 2.28 Gy, Breast = 2.12 Gy, Bladder = 2.13 Gy. Case 2 – Sequential approach: 2 fractions × 3 Gy; SIB dose = 3.3 Gy; Most frequent global decision: SIB (mode = 2)

**Figure A2:**
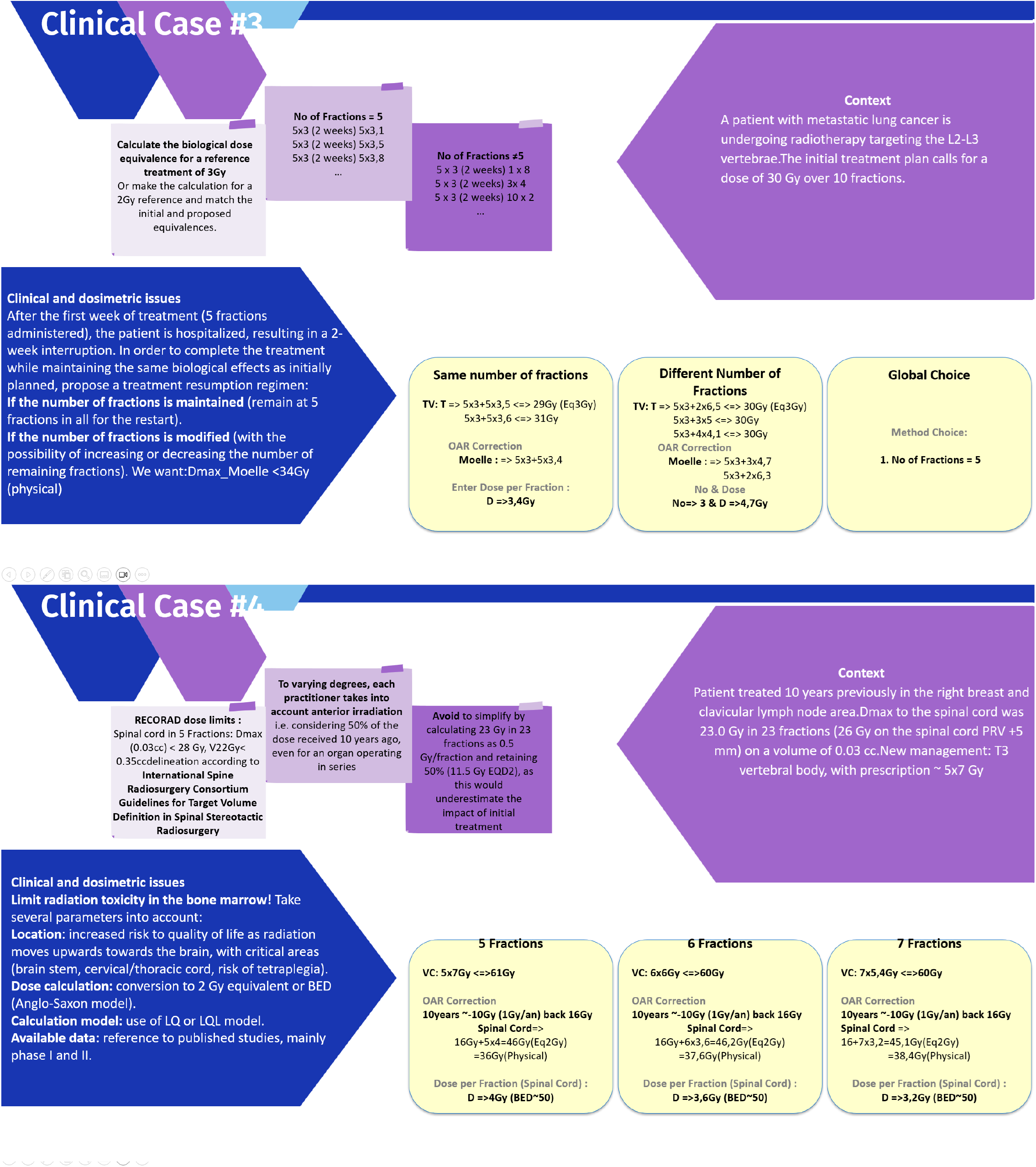
Illustration of Case Studies 3–4: Clinical Context and Example Treatment Strategies. Case 3 – Same number of fractions: 3.4 Gy; Different number: 3 × 4.7 Gy; Most frequent global decision: same number of fractions (mode = 1). Case 4 – Dose per fraction proposed by authors: 5 fractions = 4.0 Gy, 6 fractions = 3.6 Gy, 7 fractions = 3.2 Gy.

## Footnotes

1 https://www.mdapp.co/biologically-effective-dose-bed-calculator-493/, https://www.mdcalc.com/calc/10111/radiation-biologically-effective-dose-bed-calculator, and https://www.sfjro.fr/ilq/fr/ilq.html?checkbox=1

